# Remnant cholesterol associated with all-cause mortality in oldest-old acute coronary syndrome patients: a cohort study

**DOI:** 10.1101/2023.09.21.23295931

**Authors:** Boning Zhou, Ling Gao, Xin Zhang, Jian Shen, Ying Li, Yang Jiao, Jian Wang, Xiaoling Hou, Yongkang Su, Zhenhong Fu

## Abstract

**Background:** Previous studies showed that remnant cholesterol(RC) partially explained the residual risk of cardiovascular disease. We tried to test the hypothesis that remnant cholesterol is a risk factor for all-cause mortality in oldest-old(>80 years old) patients with acute coronary syndrome(ACS).

**Methods:** 662 oldest-old patients with acute coronary syndrome and undergoing coronary angiography were finally included in our study. Remnant cholesterol(RC) was calculated by fasting total cholesterol(TC) minus low-density lipoprotein cholesterol(LDL-C) and high-density lipoprotein cholesterol(HDL-C). Multivariable Cox regression analysis was used to identify the association between RC and all-cause mortality. Receiver operating characteristic(ROC) curve was used to compare the discrimination capacity of RC, TC, HDL-C and LDL-C to predict all-cause mortality.

**Results:** Among the 662 oldest-old patients enrolled, 92.90% accepted statin therapy. The average age was 81.87 ± 2.14 years old. In the fully-adjusted Cox regression model, the Hazard ratio(HR) [95% confidence interval (CI)] of all-cause mortality was 1.17 (1.01, 1.36) for each standard deviation(SD) increase in RC. Compared to tertile 1, tertile 3 was associated with a 130% increased risk of all-cause mortality (HR:2.30;95%CI:1.45, 3.63). The increased risk of death from tertile 1 to tertile 3 was statistically significant (P for trend<0.001). The subgroup analysis confirmed the significant association between RC and all-cause mortality. ROC curve showed that RC had a better discrimination capacity at predicting all-cause mortality and improved the prognostic value of the Gensini score combined with left ventricular ejection fraction(LVEF).

**Conclusion:** Remnant cholesterol was associated with all-cause mortality in oldest-old patients with ACS in a long-term follow-up.

**Key learning points:** *What is already known:*

(1) Remnant cholesterol was a risk factor for atherosclerotic cardiovascular disease(ASCVD);
(2) Emerging studies reported that RC was associated with all-cause mortality in general population, patients with disease status including ischemic heart disease(IHD), heart failure(HF) and metabolic dysfunction-associated fatty liver disease(MAFLD) or patients receiving special treatment such as peritoneal dialysis(PD) and kidney transplantation;
(3) However, some studies found that there was no correlation between RC and all-cause mortality.

*What this study adds:*

(1) We focused on oldest-old ACS patients who underwent angiography and found that RC was associated with all-cause mortality in this special population;
(2) The correlation between RC and all-cause mortality was dose–response;
(3) RC had a better discrimination capacity at predicting all-cause mortality and improved the prognostic value of the Gensini score combined with LVEF.

## Background

With the exacerbation of population aging, the proportion of elderly patients is increasing.^1^ The oldest-old people(aged 80 years old or above), an important part of elderly people, are vulnerable to stressors and result in an increase of adverse events.^2^ Coronary heart disease(CHD) is also a common chronic disease that does harm to the elderly’s health.^3^ According to the latest data, cardiovascular disease remains the leading cause of human death worldwide.^4^ Acute coronary syndrome(ACS), a severe clinical subtype of CHD, leads to the increased risk of adverse cardiovascular outcomes and all-cause mortality, especially in the oldest-old people.^5^

Remnant cholesterol(RC), the cholesterol content of triglyceride-rich lipoproteins(TGRLs), is supposed to be related with residual cardiovascular risk.^6–8^ A growing amount of evidence suggests that among statin-treated patients with high triglycerides, despite a low level of low-density lipoprotein cholesterol(LDL-C), significant residual cardiovascular risk remains.^9–11^ Triglycerides are easily metabolized while cholesterol is able to accumulate in the intima of coronary artery, be taken up by macrophages or smooth muscle cells and eventually lead to coronary atherosclerosis.^6,12,13^ So RC may play a partial role in atherosclerosis development.

Previous studies suggested that remnant cholesterol was a risk factor for cardiovascular disease^6–9,14–19^ Also, some evidence showed that there might be an association between increased remnant cholesterol and all-cause mortality in patients with or without ischemic heart disease.^20–28^ Nevertheless, few studies focused on, in the oldest-old people with ACS, the correlation between RC and all-cause mortality and comparison of the discrimination capacity of RC and LDL-C to predict all-cause mortality. Accordingly, our study aims to identify whether the increased remnant cholesterol is a risk factor for all-cause mortality in oldest-old patients with ACS in a long-term follow-up.

## Methods

### Study Participants

A total of 720 patients were enrolled. 699 patients signed the informed consent and were included in the study. During the long-term follow-up, 37 were lost, and 662 were finally available for our statistical analysis. The follow-up rate was 94.7%. To be included in the analysis, subjects were required to (1)be over 80 years old, (2)be diagnosed as acute coronary syndrome, and (3)accept coronary angiography in the Cardiology Department of Chinese PLA General Hospital from January 2006 to December 2012. The study was based on the following exclusion criteria: patients (1) with severe valvular heart disease, pulmonary hypertension, severe liver insufficiency, rheumatoid arthritis, malignant tumors, and infectious diseases; (2)with neuropsychiatric disorders that prevented them from cooperating with the researcher. Our study was performed in line with the Declaration of Helsinki and was approved by Ethics Service Center of Chinese People’s Liberation Army(PLA) General Hospital.

### Diagnose and treatment

The diagnosis of coronary heart disease and the severity of coronary artery lesions were determined by coronary angiography which was analyzed by the same image analysis software. The Gensini score was used to represent the severity of coronary artery stenosis. Being performed in the cardiac interventional center of Chinese PLA general hospital, coronary intervention and perioperative management were in line with current guidelines. According to guideline, loading doses of aspirin (300 mg) and clopidogrel (300 mg) were given before the intervention. On the basis of the results of coronary angiography and patient’s own will, patients were given individualized interventions, including intensive drug treatment, percutaneous coronary intervention (PCI) or coronary artery bypass grafting (CABG). After discharge the long-term follow-up observation was conducted.

### Data collection and definitions

All data were collected by the experimental data recorders who passed uniform professional training.

The baseline information is as follows:

(1) general information(age, gender, body mass index(BMI), heart rate(HR), systolic blood pressure(SBP), diastolic blood pressure (DBP), left ventricular ejection fraction(LVEF), and Gensini score)
(2) cardiovascular risk factors(hypertension, diabetes, hyperlipidemia, prior acute myocardial infarction(prior-AMI), previous stroke, chronic kidney injury(CKD) and current smoking)
(3) fasting blood biochemical indicators at 6 am the next day after admission(total cholesterol(TC), triglyceride(TG), low-density lipoprotein-cholesterol(LDL-C), high-density lipoprotein-cholesterol(HDL-C), Standardized creatinine(Scr), the estimated glomerular filtration rate(eGFR), fasting blood glucose(FBG), uric acid(UA))
(4) cardiovascular medication experience(aspirin, clopidogrel, statins, β-blockers(BB), angiotensin-converting enzyme inhibitor(ACEI)/angiotensin receptor blocker(ARB))
(5) culprit artery(left anterior descending artery(LAD), left circumflex artery(LCX), left main coronary artery(LM), right coronary artery(RCA), and multi-vessel lesion)
(6) treatment strategies (intensive treatment with medicine, PCI and CABG).

The remnant cholesterol was defined as fasting total cholesterol(TC) minus low-density lipoprotein cholesterol(LDL-C) minus high-density lipoprotein cholesterol(HDL-C), which is called calculated remnant cholesterol.

The body mass index(BMI) was defined as the body weight (kg) divided by height square(m^2^). The estimated glomerular filtration rate(eGFR) was calculated by the Chinese modified Modification of Diet in Renal Disease equation: eGFR(mL/min/1.73m^2^)=175×standardized creatinine(mg/dL)^−1.234^×age(year)^−0.179^×0.79 (if female).

eGFR<60 mL/min/1.73 m^2^ was considered a diagnostic indicator of Chronic Kidney Disease. Standardized creatinine(Scr) was calculated by the calibration equation: Scr(mg/dL)=0.795×enzymatic method Scr(mg/dL)+ 0.29.

According to the results of coronary angiography, more than two culprit vessels with diameter stenosis≥50% was considered coronary multivessel lesion.

### Outcome assessment

The primary endpoint was all-cause mortality. Being performed every 12 months after hospital discharge, the clinical follow-up lasted up to 10 years. The follow-up information was obtained from telephone record, family interview or corresponding medical records.

### Statical analysis

According to remnant cholesterol(RC) tertile, the participants were divided into three parts. Each One in three study population constituted a group. The normality of continuous variables was assessed by kurtosis and skewness measures and normal probability plots. The continuous variable with non-normal distribution was shown as medians with interquartile range. However, if the continuous variable was normally distributed, it would be expressed as means±standard deviation(SD). If the continuous variances were homogeneous, the t-test or analysis of variance(ANOVA) was appropriate; if the variances were not homogeneous, the rank-sum test was used. According to enumeration data, the differences between groups were assessed using the chi-square test. RC and clinical parameters were continuous variables, so Pearson’s correlation test was suitable enough to assess the associations between RC and clinical parameter. Unadjusted survival curves were generated in Kaplan–Meier plots with log-rank tests. The association between RC and all-cause mortality was estimated using Cox proportional hazards models. Three regression models were built and potential confounders were adjusted in these models. Model 1 was the unadjusted model, model 2 was the partially adjusted model that age and sex were controlled, and model 3 was the fully adjusted model that the following variables were controlled including sex, age, BMI, smoking, diabetes, hypertension, stroke, prior-AMI, hyperlipidemia, CKD, SBP, DBP, eGFR, FPG, UA, LVEF, Gensini score, aspirin, clopidogrel, statin, BB, ACEI/ARB, LM, Muti-vessel, Treatment(intensive drug treatment, percutaneous coronary intervention (PCI) or coronary artery bypass grafting (CABG)), TG, HDL-C, LDL-C and TC. ROC curve was used to compare the discrimination capacity of RC, TC, HDL-C and LDL-C to predict all-cause mortality. The basic model on the Gensini score combined with LVEF was constructed for all-cause mortality risk prediction.

The RC was transformed into three categorical variables for the primary analysis. For the trend test, the newly categorical variable was also recorded as a continuous variable and entered into the regression models. We also standardized the RC, then put it into the regression models to determine the relationships between the increase of RC per SD and Hazard ratio(HR). In addition, we conducted subgroup analysis to explore whether the association between RC and all-cause mortality was modified by the following variables: sex, hypertension, diabetes, previous stroke, prior-AMI, hyperlipidemia, CKD, current smoking, BMI and multivessel lesion. Interactions between the RC and each of the above variables were tested. Results were reported by hazard ratios (HR) and 95% confidence intervals (CI).

Two-sided P<0.05 was considered statistically significant. All data in this study were processed using SPSS software Version 23.0 (IBM Corporation, Armonk, NY, USA), the statistical software packages R (http://www.R-project.org, The R Foundation) and Empower Stats(http://www. empowerstats.com, X&Y Solutions, Inc., Boston, MA).

## Results

### Baseline characteristics of study participants

According to the tertile of RC, a total of 662 elderly patients with ACS were divided into three groups. The patients in the much higher RC group would have higher TC, TG and LDL-C and have lower HDL-C. Also, the subjects with high RC level are more likely to be female. Most importantly, in the high RC group, FBG and the percentage of patients with diabetes mellitus(DM) tended to be higher (Table 1).

**Table 1.** Baseline characteristics of participants stratified by tertile of RC.

| Variable | Total | Tertile1<br>RC < 0.36 | Tertile2<br>0.36 ≤ RC <<br>0.63 | Tertile3<br>RC ≥ 0.63 | P-value |
| --- | --- | --- | --- | --- | --- |
| N |  | 211 (31.87%) | 229 (34.59%) | 222 (33.53%) | - |
| Age | 81.87 ± 2.14 | 81.93 ± 2.28 | 81.90 ± 2.03 | 81.79 ± 2.11 | 0.758 |
| Height | 165.32 ± 8.25 | 166.41 ± 7.47 | 165.57 ± 8.92 | 164.03 ± 8.10 | **0.009** |
| Weight | 67.20 ± 10.68 | 67.18 ± 10.30 | 67.64 ± 11.13 | 66.75 ± 10.60 | 0.680 |
| BMI | 24.57 ± 3.40 | 24.22 ± 3.14 | 24.66 ± 3.59 | 24.79 ± 3.43 | 0.183 |
| HR | 74.81 ± 14.02 | 74.70 ± 14.09 | 74.06 ± 15.33 | 75.67 ± 12.47 | 0.472 |
| SBP | 137.08 ± 21.79 | 135.64 ± 20.24 | 137.79 ± 23.80 | 137.72 ± 21.05 | 0.508 |
| DBP | 71.44 ± 12.12 | 70.77 ± 11.42 | 71.90 ± 12.52 | 71.60 ± 12.36 | 0.604 |
| TC | 3.92 ± 1.12 | 3.40 ± 0.80 | 3.86 ± 0.84 | 4.49 ± 1.35 | **<0.001** |
| TG | 1.39 ± 1.00 | 0.89 ± 0.32 | 1.22 ± 0.40 | 2.04 ± 1.43 | **<0.001** |
| LDL-C | 2.18 ± 0.77 | 1.93 ± 0.73 | 2.25 ± 0.74 | 2.35 ± 0.78 | **<0.001** |
| HDL-C | 1.15 ± 0.34 | 1.32 ± 0.35 | 1.13 ± 0.28 | 1.01 ± 0.32 | **<0.001** |
| **eGFR** | 70.85 ± 23.11 | 73.60 ± 18.28 | 70.89 ± 19.32 | 68.21 ± 29.66 | 0.053 |
| **FBG** | 6.02(5.04-7.88) | 5.75 (4.85-7.47) | 5.98 (5.04-7.66) | 6.36 (5.13-8.70) | **0.002** |
| **UA** | 351.59 ± 149.66 | 338.99 ± 102.02 | 354.10 ± 198.86 | 360.99 ± 126.73 | 0.296 |
| **LVEF** | 55.65 ± 9.91 | 55.38 ± 9.30 | 56.93 ± 9.23 | 54.60 ± 10.99 | 0.143 |
| **Gensini score** | 44.00(20.00-72.00) | 48.00<br>(20.00-80.00) | 44.00<br>(20.00-68.00) | 44.00<br>(20.00-83.00) | 0.775 |
| **Female** | 186 (28.10%) | 39 (18.48%) | 55 (24.02%) | 92 (41.44%) | **<0.001** |
| **Diabetes** | 231 (34.89%) | 53 (25.12%) | 80 (34.93%) | 98 (44.14%) | **<0.001** |
| **Hypertension** | 511 (77.19%) | 166 (78.67%) | 166 (72.49%) | 179 (80.63%) | 0.099 |
| **Stroke** | 138 (20.85%) | 46 (21.80%) | 52 (22.71%) | 40 (18.02%) | 0.433 |
| **Prior-AMI** | 120 (18.13%) | 36 (17.06%) | 38 (16.59%) | 46 (20.72%) | 0.465 |
| **Hyperlipidemia** | 151 (22.81%) | 44 (20.85%) | 46 (20.09%) | 61 (27.48%) | 0.124 |
| **CKD** | 78 (11.78%) | 23 (10.90%) | 27 (11.79%) | 28 (12.61%) | 0.859 |
| **Smoking** | 164 (24.77%) | 54 (25.59%) | 62 (27.07%) | 48 (21.62%) | 0.385 |
| **Aspirin** | 642 (96.98%) | 205 (97.16%) | 222 (96.94%) | 215 (96.85%) | 0.982 |
| **Clopidogrel** | 632 (95.47%) | 200 (94.79%) | 219 (95.63%) | 213 (95.95%) | 0.836 |
| **Statin** | 615 (92.90%) | 200 (94.79%) | 217 (94.76%) | 198 (89.19%) | **0.031** |
| **BB** | 418 (63.14%) | 129 (61.14%) | 137 (59.83%) | 152 (68.47%) | 0.125 |
| **ACEI/ARB** | 364 (54.98%) | 104 (49.29%) | 132 (57.64%) | 128 (57.66%) | 0.131 |
| **LAD** | 560 (84.59%) | 175 (82.94%) | 197 (86.03%) | 188 (84.68%) | 0.668 |
| **LCX** | 383 (57.85%) | 121 (57.35%) | 133 (58.08%) | 129 (58.11%) | 0.984 |
| **RCA** | 429 (64.80%) | 135 (63.98%) | 143 (62.45%) | 151 (68.02%) | 0.443 |
| **LM** | 109 (16.47%) | 36 (17.06%) | 38 (16.59%) | 35 (15.77%) | 0.934 |
| **Multivessel** | 466 (70.39%) | 144 (68.25%) | 160 (69.87%) | 162 (72.97%) | 0.547 |
| **Treatment** |  |  |  |  | 0.461 |
| **Intensive medication** | 241 (36.40%) | 77 (36.49%) | 75 (32.75%) | 89 (40.09%) |  |
| **PCI** | 405 (61.18%) | 128 (60.66%) | 150 (65.50%) | 127 (57.21%) |  |
| **CABG** | 16 (2.42%) | 6 (2.84%) | 4 (1.75%) | 6 (2.70%) |  |

BMI, body mass index; HR heart rate; SBP, systolic blood pressure; DBP, diastolic blood pressure; TC, total cholesterol; TG, triglyceride; LDL-C, low-density lipoprotein-C; HDL-C, high-density lipoprotein-C; eGFR, estimated glomerular filtration rate; FBG, fasting blood glucose; UA, uric acid; LVEF, left ventricular ejection fraction; MI, myocardial infarction; CKD, chronic kidney disease; BB, β-blockers; ACEI, angiotensin-converting enzyme inhibitor; ARB, angiotensin receptor blocker; LAD, left anterior descending artery; LCX, left circumflex artery; RCA, right coronary artery; LM, left main coronary artery; PCI, percutaneous coronary intervention; CABG, coronary artery bypass grafting

### Correlations between RC and clinical variables

The traditional cardiovascular risk factors and RC are continuous variables and normally distributed, so Pearson’s correlation analysis was performed to identify the correlation between the RC and risk factors. As shown in Table 2, RC was negatively correlated with HDL-C and positively correlated with TC, TG, FBG, especially TC and TG. There was no association between RC and LDL-C.

**Table 2.**
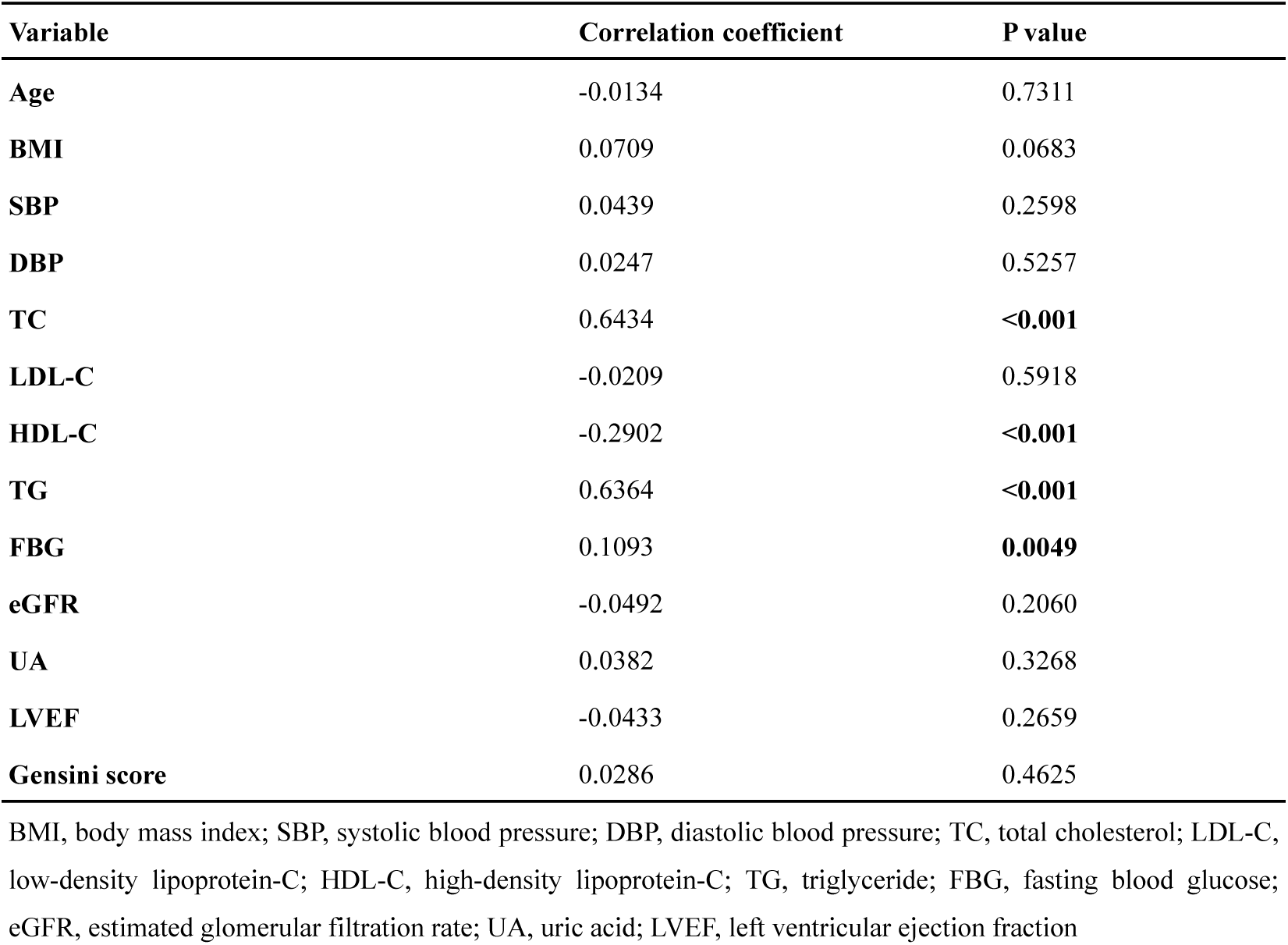
Correlations between RC and traditional cardiovascular risk factors.

| Variable | Correlation coefficient | P value |
| --- | --- | --- |
| Age | -0.0134 | 0.7311 |
| BMI | 0.0709 | 0.0683 |
| SBP | 0.0439 | 0.2598 |
| DBP | 0.0247 | 0.5257 |
| TC | 0.6434 | <0.001 |
| LDL-C | -0.0209 | 0.5918 |
| HDL-C | -0.2902 | <0.001 |
| TG | 0.6364 | <0.001 |
| FBG | 0.1093 | 0.0049 |
| eGFR | -0.0492 | 0.2060 |
| UA | 0.0382 | 0.3268 |
| LVEF | -0.0433 | 0.2659 |
| Gensini score | 0.0286 | 0.4625 |

BMI, body mass index; SBP, systolic blood pressure; DBP, diastolic blood pressure; TC, total cholesterol; LDL-C, low-density lipoprotein-C; HDL-C, high-density lipoprotein-C; TG, triglyceride; FBG, fasting blood glucose; eGFR, estimated glomerular filtration rate; UA, uric acid; LVEF, left ventricular ejection fraction

### Association between RC and all-cause mortality

As shown in Table 3, in the fully adjusted regression model(model 3), for each standard deviation increase of RC, the risk of death increased by 17%. Compared to tertile 1, tertile 3 was associated with a 130% increased risk of all-cause mortality (HR:2.30; 95%CI:1.45, 3.63). Whether it is an unadjusted model, a partially adjusted model or a fully adjusted model, the increased risk of death from tertile 1 to tertile 3 was statistically significant (P for trend<0.001). As presented in Figure 2, the Kaplan Meier curves of the long-term survival in oldest-old ACS patients showed that the survival probability in tertile 3 was significantly lower than that in tertile 1.(P < 0.0001).

**Figure 1.**
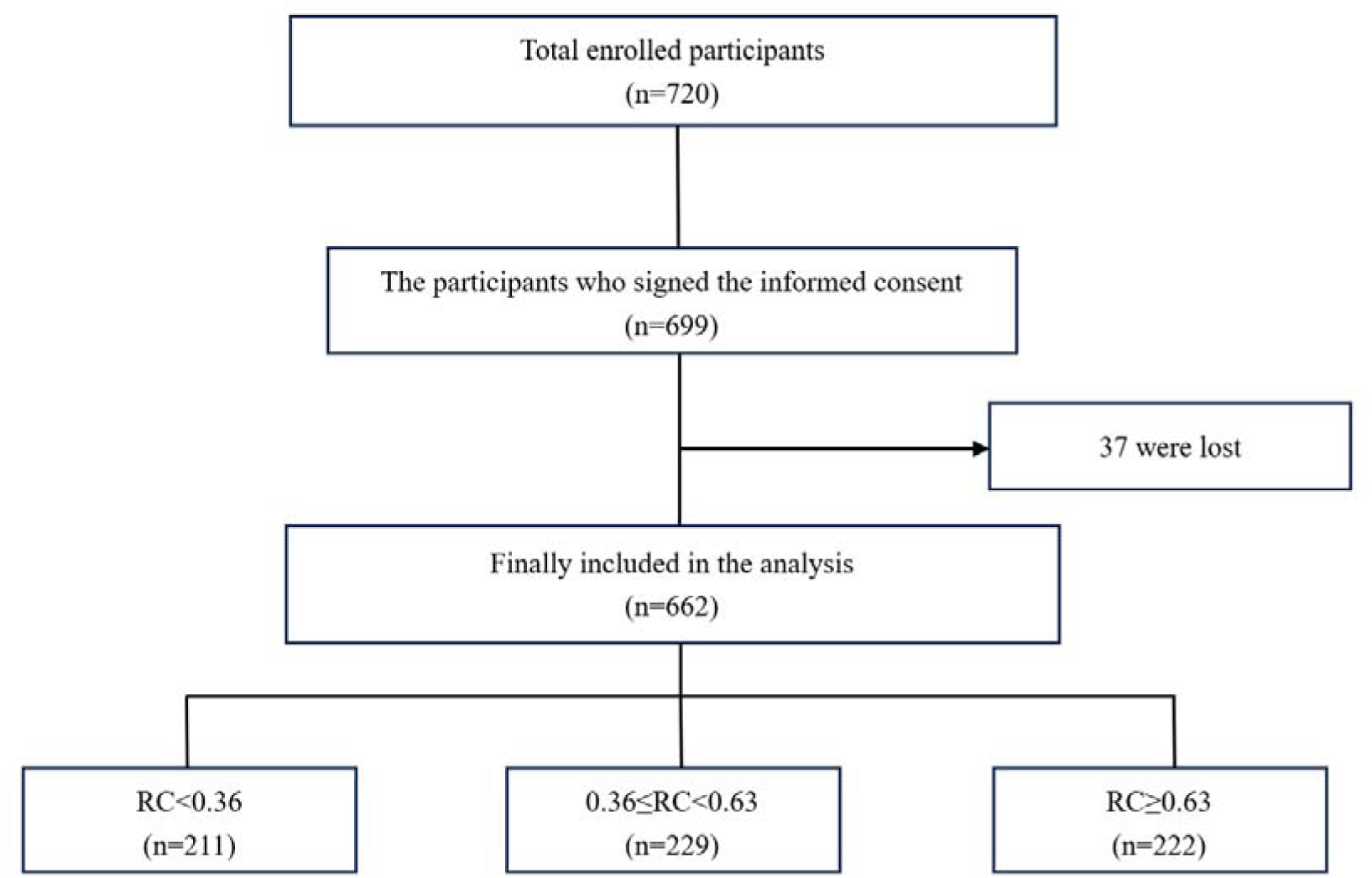
Flow chart of the study participants.

**Table 3.**
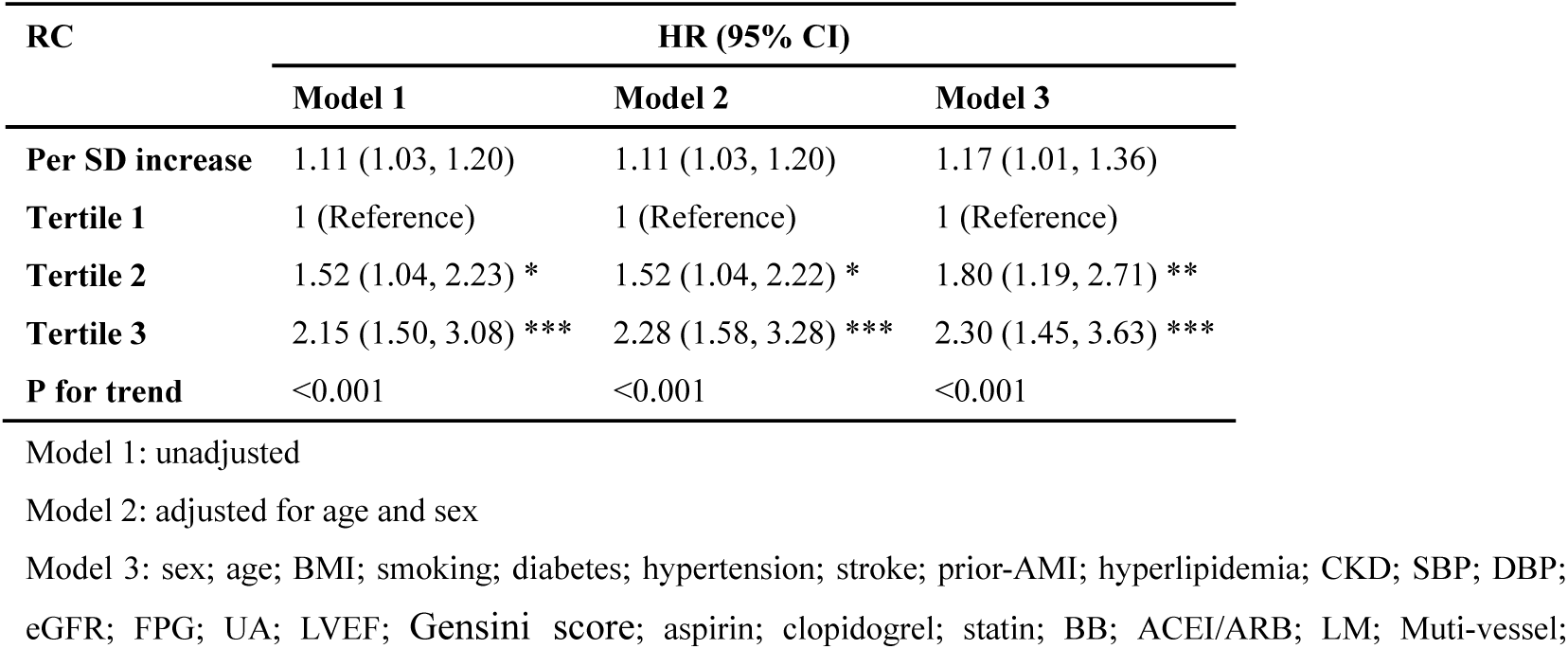
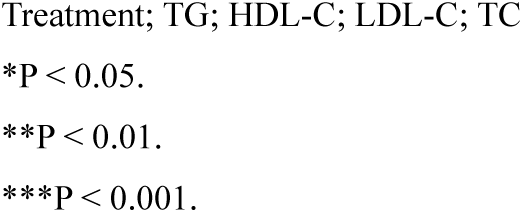
Multivariable Cox regression analysis for the correlation between RC and all-cause mortality.

**Figure 2.**
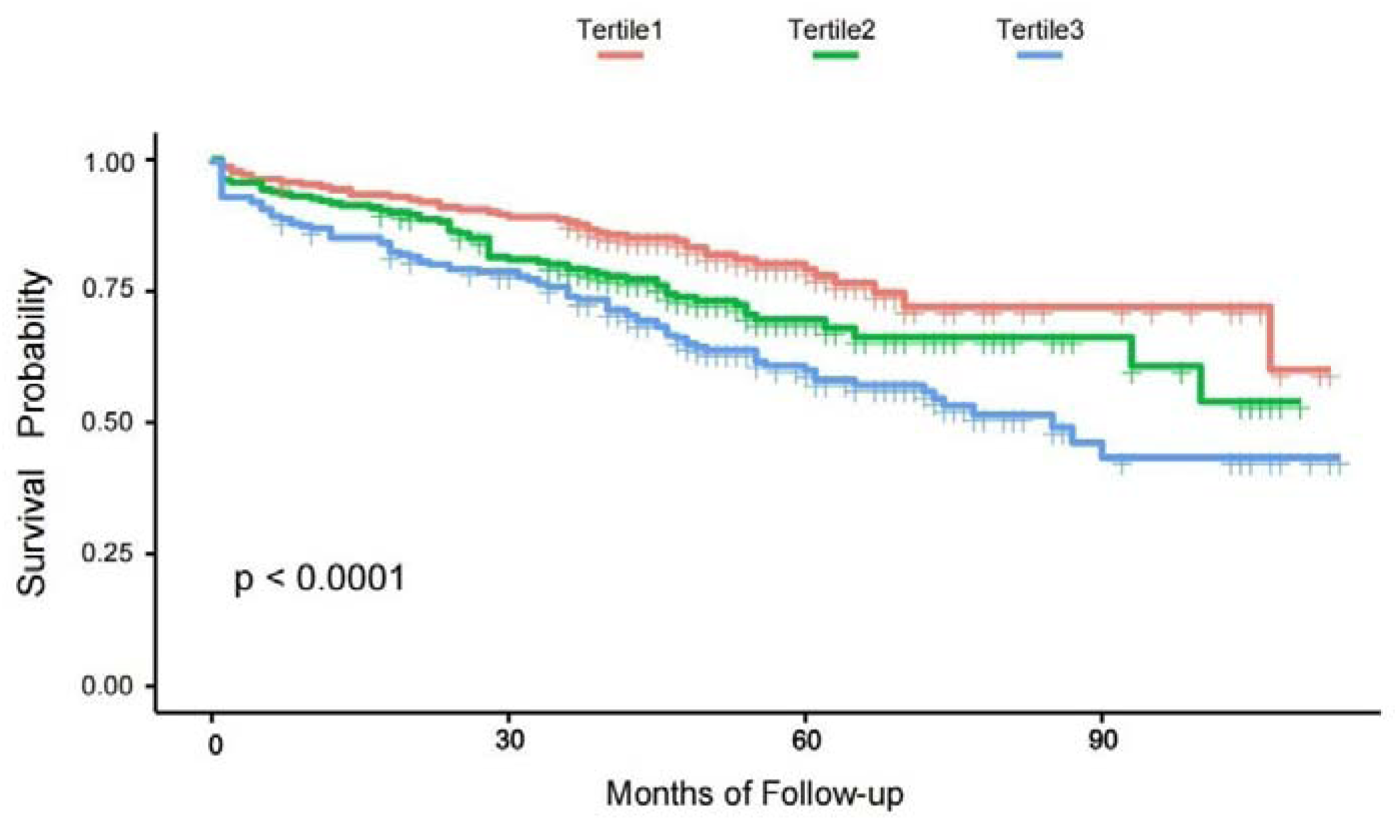
Kaplan–Meier survival curves for the association between RC and all-cause mortality

### Subgroup analysis for correlation between RC and long-term prognosis

To analyze the interaction between RC and other variables, subgroup analysis was performed, further exploring the correlation between RC and all-cause mortality. As shown in Figure 3, in subgroup analysis, there was no significant interaction between RC and the following variable, including sex, DM, Hypertension, stroke, CKD and LDL-C.

**Figure 3.**
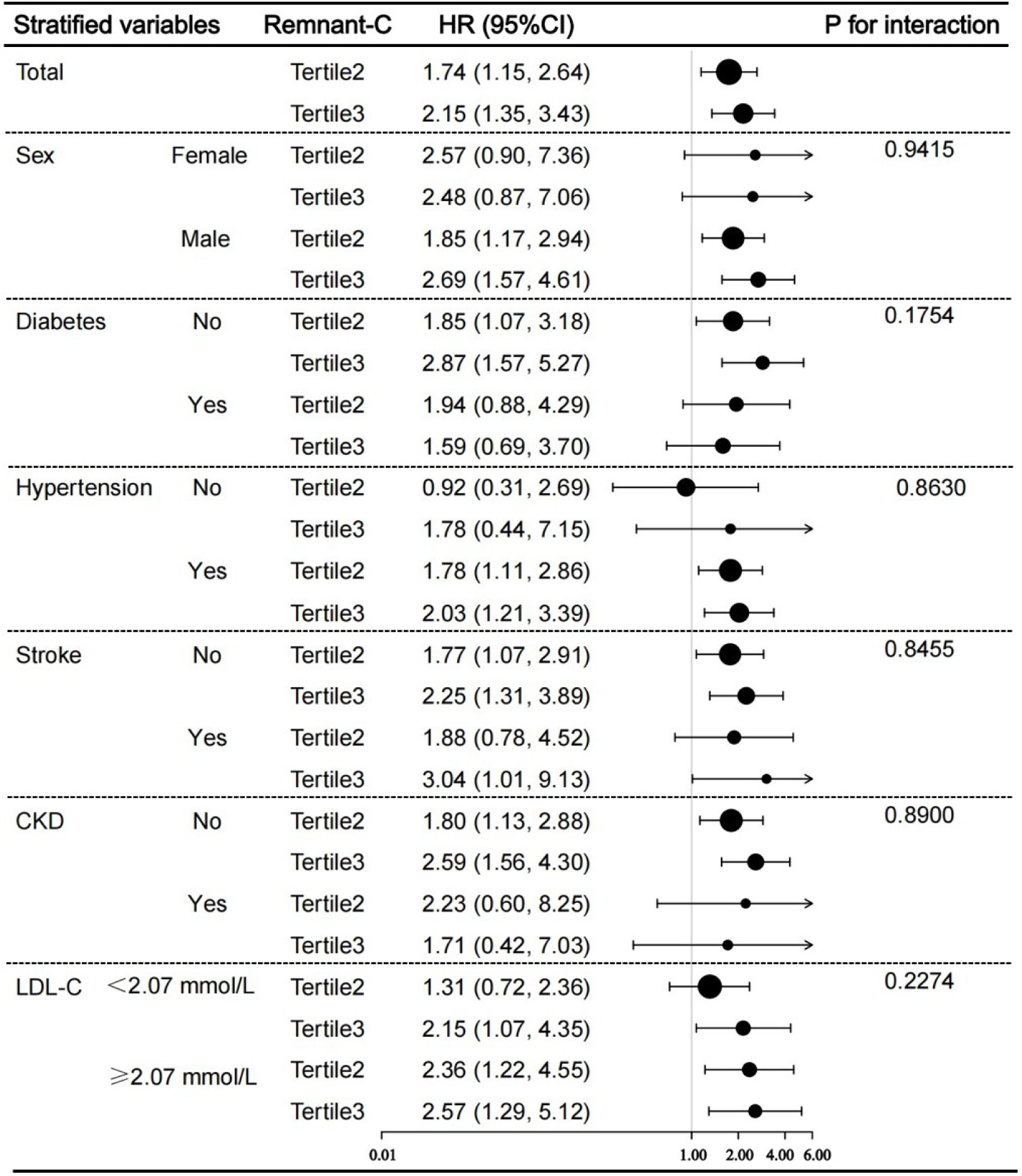
Forest plot investigating the association between RC and all-cause mortality in different subgroups. Adjusted for sex; age; BMI; smoking; diabetes; hypertension; stroke; prior-AMI; hyperlipidemia; CKD; SBP; DBP; eGFR; FPG; UA; LVEF; Gensini score; aspirin; clopidogrel; statin; BB; ACEI/ARB; LM; Muti-vessel; Treatment; TG; HDL-C; LDL-C; TC except for the stratified variable.

### Comparisons of the capacity of RC, TC, HDL-C and LDL-C to predict all-cause mortality

To predict all-cause mortality, the discrimination capacity of RC, TC, HDL-C and LDL-C was compared by ROC curve. RC has slightly less sensitivity than LDL-C and TC, but has higher sensitivity than HDL-C. The specificity of RC outperformed LDL-C and TC, but slightly underperformed HDL-C. More importantly, RC had the highest Youden’s index, accuracy, positive predictive value and negative predictive value (Figure 4a). The AUC of the RC, TC, HDL-C and LDL-C was 0.621 (0.575-0.666), 0.549 (0.502-0.597), 0.582 (0.534-0.630) and 0.515 (0.467-0.564) respectively(Figure 4b). As shown in Table 4, RC was superior to LDL-C at predicting all-cause mortality according to the discrimination index values including AUC, NRI and IDI. To further examine the ability of RC, TC, HDL-C, and LDL-C at predicting all-cause mortality, the basic model on the Gensini score combined with LVEF was constructed for all-cause mortality risk prediction. The AUC of the Gensini score + LVEF, the Gensini score + LVEF + RC, the Gensini score + LVEF + TC, the Gensini score + LVEF + HDL-C, the Gensini score + LVEF + LDL-C was 0.625, 0.654 (P<0.001), 0.630 (P=0.449), 0.643(P=0.117), 0.625(P=0.642), respectively(Figure 4c). We found that only RC provided a significant difference in AUC(P<0.001) and improved the prognostic value of the Gensini score combined with LVEF.

**Table 4.**
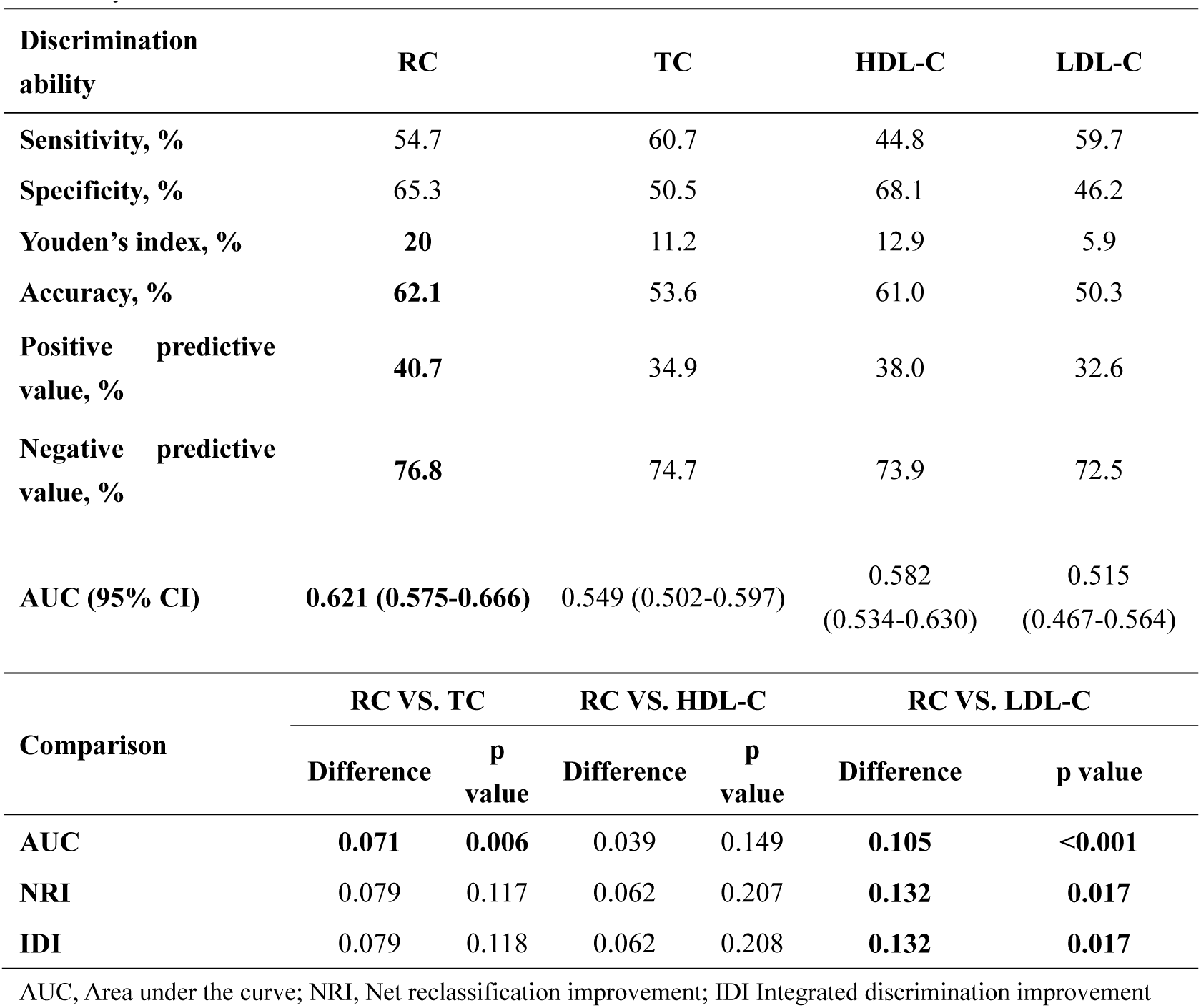
Comparative analysis of the discrimination of RC, TC, HDL-C and LDL-C for all-cause mortality.

**Table 5.** Model performance after the addition of RC, TC, HDL-C or LDL-C to the basic model (Gensini score and LVEF) for predicting all-cause mortality.

| **Model** | **AUC** | **P value** | **NRI** | **P value** | **IDI** | **P value** |
| --- | --- | --- | --- | --- | --- | --- |
| **Gensini score + LVEF** | 0.625 | - | - | - | - | - |
| **Gensini score + LVEF + RC** | **0.654** | **<0.001** | 0.014 | 0.644 | 0.014 | 0.645 |
| **Gensini score + LVEF + TC** | 0.630 | 0.449 | 0.010 | 0.542 | 0.010 | 0.542 |
| Gensini score + LVEF + HDL-C | 0.643 | 0.117 | 0.017 | 0.565 | 0.017 | 0.566 |
| Gensini score + LVEF + LDL-C | 0.625 | 0.642 | 0.007 | 0.535 | 0.007 | 0.536 |

**Figure 4.**
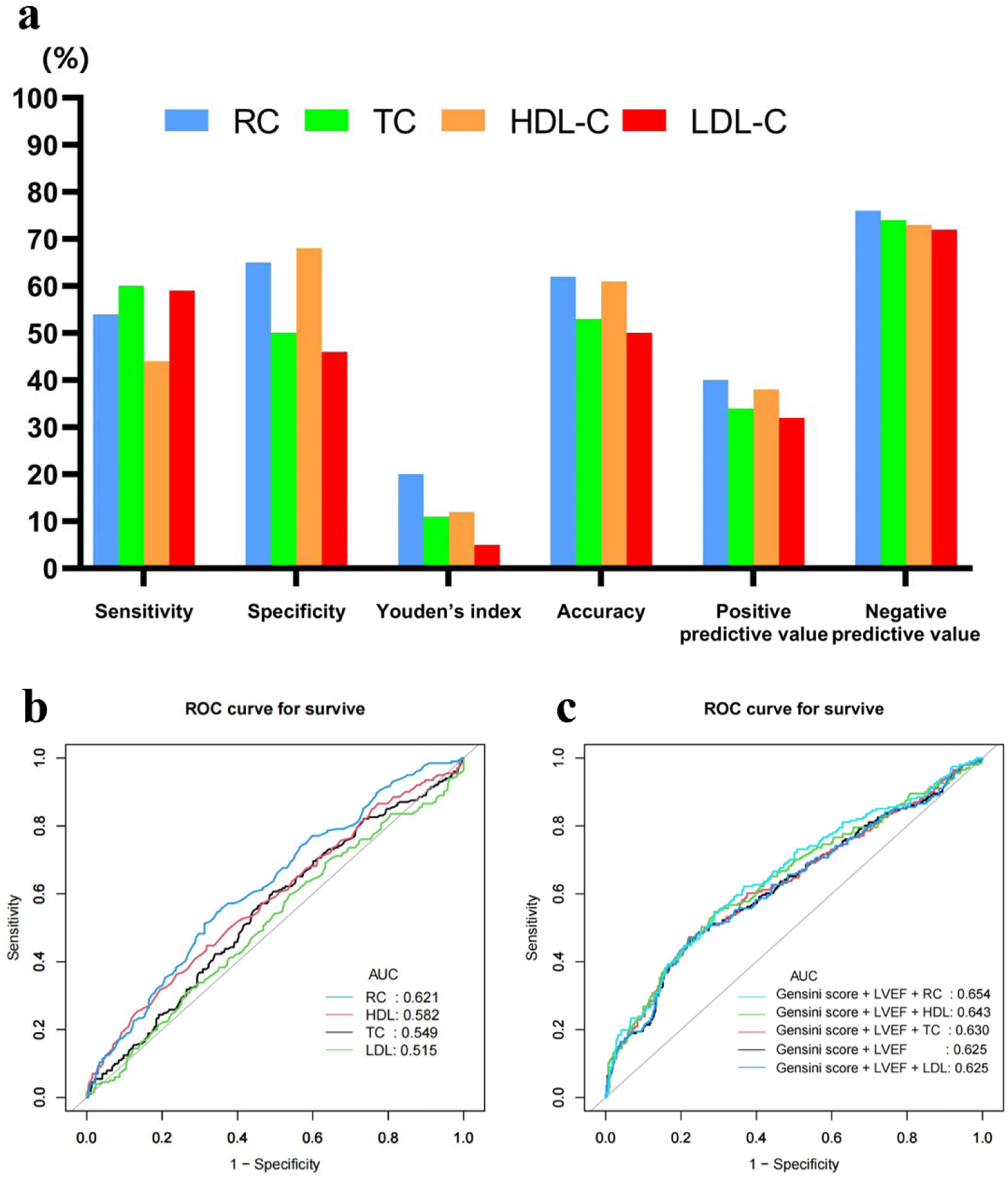
a. Diagnostic performance of RC, TC, HDL-C and LDL-C to predict all-cause mortality; **b** AUC of RC, TC, HDL-C and LDL-C at predicting all-cause mortality; **c** AUC of the basic model(Gensini score and LVEF), +RC, +TC, +HDL-C or +LDL-C for prediction of all-cause mortality

## Discussion

This study focused on 662 ACS patients(>80 years old) who underwent coronary angiography and accepted about 10 years follow-up. As far as we know, this study, for the first time, established the correlation between the plasm remnant cholesterol (RC) level and all-cause mortality in the oldest-old people with ACS. The main findings of our study were as follows: (1) The significant association between RC and all-cause mortality was established whether RC was regarded as a continuous or categorical variable. We also found that the correlation between RC and all-cause mortality was dose–response; (2) RC had a better discrimination capacity at predicting all-cause mortality and improved the prognostic value of the Gensini score combined with LVEF.

Several studies reported that there is a potential association between RC and all-cause mortality in general population^20–22^, patients with disease status including ischemic heart disease(IHD)^23^, heart failure(HF)^24^and metabolic dysfunction-associated fatty liver disease(MAFLD)^25^ or patients receiving special treatment such as peritoneal dialysis(PD)^26^ and kidney transplantation^27^. Besides, one meta-analysis also reported that RC is associated with all-cause mortality.^28^ However, some emerging evidence found a conflicting conclusion on the relationship between RC and all-cause mortality. Gao S et al reported that there is no association between RC and all-cause mortality in patients with myocardial infarction with nonobstructive coronary arteries(MINOCA)^29^. Moreover, Tian Y et al reviewed published articles and found that RC is not associated with all-cause mortality.^30^ Therefore, the relationship between RC and all-cause mortality remained inconclusive. Considering the aging population, as well as the huge burden on medical and health care work brought by cardiovascular disease, it is high time we should explore the relationship between RC and all-cause mortality in oldest-old ACS people.

So far, no studies focused on oldest-old ACS patients and figured out whether the association between RC and all-cause mortality exists in this special population. According to ACS patients, there are merely two studies that attempted to establish the association between RC and all-cause mortality.^31,32^ Unfortunately, the results were at opposite poles. The discrepancies within these two studies were likely attributed to variations in the study population characteristics(age, the percent of DM, statin use and PCI etc.), time of follow-up, and adjustment for confounders. There were also differences between our study and two previous studies. Compared to previous studies focusing on patients around 60 years old, our study aimed at the oldest-old people(aged 81.87 ± 2.14). Moreover, the percentage of subjects who accepted statin treatment in two previous studies were only 28.69% and 71.7%, respectively.^31,32^ In our study, 92.90% study participants accepted statin therapy, suggesting that the association between RC and all-cause mortality was achieved with control for LDL-C. In order to improve the reliability and validity of our study, we adopted a variety of statistical analysis strategies, including correlation analysis, multi-variable type verification(continuous and categorical variable types), trend testing, subgroup analysis, interaction analysis. Fortunately, the correlation between RC and all-cause mortality were stable and significant in different statistical analysis models. In addition, previous studies mainly used COX regression analysis to test the relationship between RC and all-cause mortality, without further subgroup analysis and ROC curve. We further compared the discrimination capacity of RC and LDL-C to predict all-cause mortality and found that RC had a much higher Youden’s index(20 vs 5.9), specificity(65.3 vs 46.2), accuracy(62.1 vs 50.3), positive predictive value(40.7 vs 32.6), negative predictive value(76.8 vs 72.5) and AUC(0.621 vs 0.515) than LDL-C, which indicated a better discrimination capacity of RC. Furthermore, these results suggested that on the premise that LDL-C was controlled, effective management of RC could further reduce all-cause mortality in this population. In clinical practice, while focusing on LDL-C, attention should also be paid to the level of RC control. The management of RC could further improve the long-term prognosis of elderly patients with ACS, and would be a worthful supplement to LDL-C in the prognostic evaluation system.

The possible mechanisms by which elevated RC could lead to an increase of all-cause mortality in oldest-old ACS patients are as follows. Firstly, A recent study provided evidence that RC was associated with diabetes mellitus(DM), which was a significant risk factor for all-cause mortality.^33–35^ We suspected that RC could induce insulin resistance and pro-inflammatory state, leading to the occurrence of DM and increased risk of all-cause mortality. In our study, we found that fasting blood glucose(FBG) and the incidence of DM in the high RC group was significantly higher than that in the low RC group, which was also consistent with this mechanism. Secondly, triglyceride-rich lipoproteins(TGRLs) are the carriers of triglycerides and remnant cholesterol. New insight strongly suggested that elevated TGRLs represented causal risk factors for all-cause mortality.^36^ Previous studies also showed that high triglycerides were strongly associated with all-cause mortality.^9,10,37^ Nevertheless, triglycerides can be easily metabolized in most cells. Therefore, it is reasonable for us to assume that TG was just a marker of remnant cholesterol and remnant cholesterol, not triglycerides, was the harmful component in TGRLs as well as the real cause of increased all-cause mortality. In our study, we found a positive correlation between RC and TG through Pearson correlation analysis, which further verified that the relationship between TG and all-cause mortality may be caused by RC. Thirdly, RC was able to accumulate in the tunica intima of coronary artery, be taken up by macrophages or smooth muscle cells through apolipoprotein E without oxidative modification and eventually lead to formation of foam cell formation and development of atherosclerotic plaque.^7,12,13^ Besides, RC was involved in the inflammatory process of coronary atherosclerosis.^38^ Also, RC could activate platelets, resulting in platelet aggregation, activation of the coagulation cascade and formation of atherothrombosis.^9,39^ Inflammation, progression of atherosclerotic plaques and formation of atherothrombosis would exacerbate the symptoms of patients with ACS whose prognosis are already poor, and result in more major adverse cardiovascular events. Mounting evidence indicated that RC promoted coronary atherosclerosis, coronary artery calcium(CAC) progression, atherosclerotic cardiovascular disease (ASCVD) and even led to cardiovascular death.^7,9,40–44^. Regrettably, in our study, no association was found between RC and Gensini score, which represented the severity of the coronary lesion. Considering that coronary artery lesions are generally serious in elderly patients, it is hoped that a better comprehensive evaluation system for assessing the degree of stenosis and coronary calcification score could be applied in the future so as to better predict the prognosis of elderly patients with ACS. In addition to cardiovascular death, some non-cardiovascular diseases related to increased mortality, such as metabolic dysfunction-associated fatty liver, were associated with elevated RC concentrations, which might be another reason why RC affected all-cause mortality.^45,46^

### Strengths and limitations

In terms of strengths, our study is the first time to choose oldest-old ACS patients as subjects and established the correlation between RC and all-cause mortality. Also, we found that RC had a better discrimination capacity at predicting all-cause mortality. However, there are still some limitations in our study. Firstly, it is a single-center study with relatively small sample size. So further multi-center and large-sample studies are needed to verify the correlation between RC and all-cause mortality. Secondly, despite our control for many confounding factors in the analysis, some known factors (dietary habits and nutritional status such as albumin and pro-albumin) or unknown factors may be ignored. Thirdly, directly measured remnant cholesterol is superior to calculated remnant cholesterol. ^47^ So using directly measured remnant cholesterol to analyze the correlation between RC and all-cause mortality may be a more effective way. Fourthly, fasting RC occurs only a few hours before breakfast while non-fasting RC occurs in most of the 24-hour period and better represents the atherogenic lipoprotein levels. So non-fasting RC may be a better predictor of cardiovascular risk and all-cause mortality.^48–50^

## Conclusion

Remnant cholesterol was associated with all-cause mortality in oldest-old patients with ACS in a long-term follow-up. On the premise that LDL-C target was achieved, RC was a worthful supplement in the prognostic evaluation system of this population. Moreover, large randomized controlled trials(RCTs) are warranted to confirm the clinical impact of remnant cholesterol elevation on all-cause mortality in this population.

## Acknowledgements

We appreciate the study participants for the information they provided and all the staff involved in this study. The authors’ responsibilities were as follows: BNZ: data interpretation, and the first draft of the manuscript writing; LG and XZ: statistical analysis; JS, YL, YJ and JHW: data collection; ZHF, YKS and XLH: manuscript revision; YKS and ZHF: study design. All authors: searched the literature, read and approved the final manuscript.

## Data availability

The datasets used and/or analyzed during the current study are available from the corresponding author on reasonable request.

## Ethics approval and consent to participate

The study was performed in accordance with the Declaration of Helsinki and approved by the medical ethics committee of the Chinese PLA General Hospital. All participants provided written informed consent.

## References

1. Partridge L, Deelen J, Slagboom PE. Facing up to the global challenges of ageing. Nature. 2018;561:45–56. doi: 10.1038/s41586-018-0457-8

2. Clegg A, Young J, Iliffe S, Rikkert MO, Rockwood K. Frailty in elderly people. Lancet (London, England). 2013;381:752–762. doi: 10.1016/s0140-6736(12)62167-9

3. Fang EF, Xie C, Schenkel JA, Wu C, Long Q, Cui H, Aman Y, Frank J, Liao J, Zou H, et al. A research agenda for ageing in China in the 21st century (2nd edition): Focusing on basic and translational research, long-term care, policy and social networks. Ageing research reviews. 2020;64:101174. doi: 10.1016/j.arr.2020.101174

4. Tsao CW, Aday AW, Almarzooq ZI, Alonso A, Beaton AZ, Bittencourt MS, Boehme AK, Buxton AE, Carson AP, Commodore-Mensah Y, et al. Heart Disease and Stroke Statistics-2022 Update: A Report From the American Heart Association. Circulation. 2022;145:e153–e639. doi: 10.1161/CIR.0000000000001052

5. Man C, Xiang S, Fan Y. Frailty for predicting all-cause mortality in elderly acute coronary syndrome patients: A meta-analysis. Ageing research reviews. 2019;52:1–6. doi: 10.1016/j.arr.2019.03.003

6. Castañer O, Pintó X, Subirana I, Amor AJ, Ros E, Hernáez Á, Martínez-González M, Corella D, Salas-Salvadó J, Estruch R, et al. Remnant Cholesterol, Not LDL Cholesterol, Is Associated With Incident Cardiovascular Disease. Journal of the American College of Cardiology. 2020;76:2712–2724. doi: 10.1016/j.jacc.2020.10.008

7. Quispe R, Martin SS, Michos ED, Lamba I, Blumenthal RS, Saeed A, Lima J, Puri R, Nomura S, Tsai M, et al. Remnant cholesterol predicts cardiovascular disease beyond LDL and ApoB: a primary prevention study. European heart journal. 2021;42:4324–4332. doi: 10.1093/eurheartj/ehab432

8. Wilson PWF, Remaley AT. Ischemic Heart Disease Risk and Remnant Cholesterol Levels. Journal of the American College of Cardiology. 2022;79:2398–2400. doi: 10.1016/j.jacc.2022.04.016

9. Sandesara PB, Virani SS, Fazio S, Shapiro MD. The Forgotten Lipids: Triglycerides, Remnant Cholesterol, and Atherosclerotic Cardiovascular Disease Risk. Endocrine reviews. 2019;40:537–557. doi: 10.1210/er.2018-00184

10. Nordestgaard BG, Varbo A. Triglycerides and cardiovascular disease. Lancet (London, England). 2014;384:626–635. doi: 10.1016/s0140-6736(14)61177-6

11. Schwartz GG, Abt M, Bao W, DeMicco D, Kallend D, Miller M, Mundl H, Olsson AG. Fasting triglycerides predict recurrent ischemic events in patients with acute coronary syndrome treated with statins. Journal of the American College of Cardiology. 2015;65:2267–2275. doi: 10.1016/j.jacc.2015.03.544

12. Nordestgaard BG, Tybjaerg-Hansen A, Lewis B. Influx in vivo of low density, intermediate density, and very low density lipoproteins into aortic intimas of genetically hyperlipidemic rabbits. Roles of plasma concentrations, extent of aortic lesion, and lipoprotein particle size as determinants. Arterioscler Thromb. 1992;12:6–18. doi: 10.1161/01.atv.12.1.6

13. Nordestgaard BG, Wootton R, Lewis B. Selective retention of VLDL, IDL, and LDL in the arterial intima of genetically hyperlipidemic rabbits in vivo. Molecular size as a determinant of fractional loss from the intima-inner media. Arterioscler Thromb Vasc Biol. 1995;15:534–542. doi: 10.1161/01.atv.15.4.534

14. Cully M. Lipids: Remnant cholesterol is associated with ischaemic heart disease. Nature reviews Cardiology. 2013;10:119. doi: 10.1038/nrcardio.2013.3

15. Goliasch G, Wiesbauer F, Blessberger H, Demyanets S, Wojta J, Huber K, Maurer G, Schillinger M, Speidl WS. Premature myocardial infarction is strongly associated with increased levels of remnant cholesterol. Journal of clinical lipidology. 2015;9:801–806.e801. doi: 10.1016/j.jacl.2015.08.009

16. Mearns BM. Dyslipidaemia: Role of remnant cholesterol in IHD. Nature reviews Cardiology. 2013;10:553. doi: 10.1038/nrcardio.2013.132

17. Varbo A, Benn M, Nordestgaard BG. Remnant cholesterol as a cause of ischemic heart disease: evidence, definition, measurement, atherogenicity, high risk patients, and present and future treatment. Pharmacology & therapeutics. 2014;141:358–367. doi: 10.1016/j.pharmthera.2013.11.008

18. Varbo A, Freiberg JJ, Nordestgaard BG. Remnant Cholesterol and Myocardial Infarction in Normal Weight, Overweight, and Obese Individuals from the Copenhagen General Population Study. Clin Chem. 2018;64:219–230. doi: 10.1373/clinchem.2017.279463

19. Varbo A, Nordestgaard BG. Remnant Cholesterol and Triglyceride-Rich Lipoproteins in Atherosclerosis Progression and Cardiovascular Disease. Arterioscler Thromb Vasc Biol. 2016;36:2133–2135. doi: 10.1161/atvbaha.116.308305

20. Varbo A, Freiberg JJ, Nordestgaard BG. Extreme nonfasting remnant cholesterol vs extreme LDL cholesterol as contributors to cardiovascular disease and all-cause mortality in 90000 individuals from the general population. Clin Chem. 2015;61:533–543. doi: 10.1373/clinchem.2014.234146

21. Langsted A, Freiberg JJ, Tybjaerg-Hansen A, Schnohr P, Jensen GB, Nordestgaard BG. Nonfasting cholesterol and triglycerides and association with risk of myocardial infarction and total mortality: the Copenhagen City Heart Study with 31 years of follow-up. Journal of internal medicine. 2011;270:65–75. doi: 10.1111/j.1365-2796.2010.02333.x

22. Wadström BN, Pedersen KM, Wulff AB, Nordestgaard BG. Elevated remnant cholesterol, plasma triglycerides, and cardiovascular and non-cardiovascular mortality. European heart journal. 2023;44:1432–1445. doi: 10.1093/eurheartj/ehac822

23. Jepsen AM, Langsted A, Varbo A, Bang LE, Kamstrup PR, Nordestgaard BG. Increased Remnant Cholesterol Explains Part of Residual Risk of All-Cause Mortality in 5414 Patients with Ischemic Heart Disease. Clin Chem. 2016;62:593–604. doi: 10.1373/clinchem.2015.253757

24. Zhao L, Zhao X, Tian P, Liang L, Huang B, Huang L, Feng J, Zhang Y, Zhang J. Predictive value of remnant cholesterol level for all-cause mortality in heart failure patients. Frontiers in cardiovascular medicine. 2023;10:1063562. doi: 10.3389/fcvm.2023.1063562

25. Huang H, Guo Y, Liu Z, Zeng Y, Chen Y, Xu C. Remnant Cholesterol Predicts Long-term Mortality of Patients With Metabolic Dysfunction-associated Fatty Liver Disease. The Journal of clinical endocrinology and metabolism. 2022;107:e3295–e3303. doi: 10.1210/clinem/dgac283

26. Deng J, Tang R, Chen J, Zhou Q, Zhan X, Long H, Peng F, Wang X, Wen Y, Feng X, et al. Remnant cholesterol as a risk factor for all-cause and cardiovascular mortality in incident peritoneal dialysis patients. Nutrition, metabolism, and cardiovascular diseases : NMCD. 2023;33:1049–1056. doi: 10.1016/j.numecd.2023.02.009

27. Horace RW, Roberts M, Shireman TI, Merhi B, Jacques P, Bostom AG, Liu S, Eaton CB. Remnant cholesterol is prospectively associated with cardiovascular disease events and all-cause mortality in kidney transplant recipients: the FAVORIT study. Nephrology, dialysis, transplantation : official publication of the European Dialysis and Transplant Association - European Renal Association. 2022;37:382–389. doi: 10.1093/ndt/gfab068

28. Yang XH, Zhang BL, Cheng Y, Fu SK, Jin HM. Association of remnant cholesterol with risk of cardiovascular disease events, stroke, and mortality: A systemic review and meta-analysis. Atherosclerosis. 2023;371:21–31. doi: 10.1016/j.atherosclerosis.2023.03.012

29. Gao S, Xu H, Ma W, Yuan J, Yu M. Remnant Cholesterol Predicts Risk of Cardiovascular Events in Patients With Myocardial Infarction With Nonobstructive Coronary Arteries. Journal of the American Heart Association. 2022;11:e024366. doi: 10.1161/jaha.121.024366

30. Tian Y, Wu W, Qin L, Yu X, Cai L, Wang H, Zhang Z. Prognostic value of remnant cholesterol in patients with coronary heart disease: A systematic review and meta-analysis of cohort studies. Frontiers in cardiovascular medicine. 2022;9:951523. doi: 10.3389/fcvm.2022.951523

31. Cordero A, Alvarez-Alvarez B, Escribano D, García-Acuña JM, Cid-Alvarez B, Rodríguez-Mañero M, Quintanilla MA, Agra-Bermejo R, Zuazola P, González-Juanatey JR. Remnant cholesterol in patients admitted for acute coronary syndromes. European journal of preventive cardiology. 2023;30:340–348. doi: 10.1093/eurjpc/zwac286

32. Shao Q, Yang Z, Wang Y, Li Q, Han K, Liang J, Shen H, Liu X, Zhou Y, Ma X, et al. Elevated Remnant Cholesterol is Associated with Adverse Cardiovascular Outcomes in Patients with Acute Coronary Syndrome. Journal of atherosclerosis and thrombosis. 2022;29:1808–1822. doi: 10.5551/jat.63397

33. Hu X, Liu Q, Guo X, Wang W, Yu B, Liang B, Zhou Y, Dong H, Lin J. The role of remnant cholesterol beyond low-density lipoprotein cholesterol in diabetes mellitus. Cardiovascular diabetology. 2022;21:117. doi: 10.1186/s12933-022-01554-0

34. Sinclair AJ, Robert IE, Croxson SC. Mortality in older people with diabetes mellitus. Diabetic medicine : a journal of the British Diabetic Association. 1997;14:639–647. doi: 10.1002/(sici)1096-9136(199708)14:8<639::Aid-dia433>3.0.Co;2-c

35. Htay T, Soe K, Lopez-Perez A, Doan AH, Romagosa MA, Aung K. Mortality and Cardiovascular Disease in Type 1 and Type 2 Diabetes. Current cardiology reports. 2019;21:45. doi: 10.1007/s11886-019-1133-9

36. Nordestgaard BG. Triglyceride-Rich Lipoproteins and Atherosclerotic Cardiovascular Disease: New Insights From Epidemiology, Genetics, and Biology. Circulation research. 2016;118:547–563. doi: 10.1161/circresaha.115.306249

37. Thomsen M, Varbo A, Tybjærg-Hansen A, Nordestgaard BG. Low nonfasting triglycerides and reduced all-cause mortality: a mendelian randomization study. Clin Chem. 2014;60:737–746. doi: 10.1373/clinchem.2013.219881

38. Varbo A, Benn M, Tybjærg-Hansen A, Nordestgaard BG. Elevated remnant cholesterol causes both low-grade inflammation and ischemic heart disease, whereas elevated low-density lipoprotein cholesterol causes ischemic heart disease without inflammation. Circulation. 2013;128:1298–1309. doi: 10.1161/circulationaha.113.003008

39. Olufadi R, Byrne CD. Effects of VLDL and remnant particles on platelets. Pathophysiology of haemostasis and thrombosis. 2006;35:281–291. doi: 10.1159/000093221

40. Langsted A, Madsen CM, Nordestgaard BG. Contribution of remnant cholesterol to cardiovascular risk. Journal of internal medicine. 2020;288:116–127. doi: 10.1111/joim.13059

41. Li W, Huang Z, Fang W, Wang X, Cai Z, Chen G, Wu W, Chen Z, Wu S, Chen Y. Remnant Cholesterol Variability and Incident Ischemic Stroke in the General Population. Stroke. 2022;53:1934–1941. doi: 10.1161/strokeaha.121.037756

42. Ohmura H. Contribution of Remnant Cholesterol to Coronary Atherosclerosis. Journal of atherosclerosis and thrombosis. 2022;29:1706–1708. doi: 10.5551/jat.ED205

43. Wadström BN, Wulff AB, Pedersen KM, Jensen GB, Nordestgaard BG. Elevated remnant cholesterol increases the risk of peripheral artery disease, myocardial infarction, and ischaemic stroke: a cohort-based study. European heart journal. 2022;43:3258–3269. doi: 10.1093/eurheartj/ehab705

44. Zhang K, Qi X, Zhu F, Dong Q, Gou Z, Wang F, Xiao L, Li M, Chen L, Wang Y, et al. Remnant cholesterol is associated with cardiovascular mortality. Frontiers in cardiovascular medicine. 2022;9:984711. doi: 10.3389/fcvm.2022.984711

45. Zou Y, Lan J, Zhong Y, Yang S, Zhang H, Xie G. Association of remnant cholesterol with nonalcoholic fatty liver disease: a general population-based study. Lipids in health and disease. 2021;20:139. doi: 10.1186/s12944-021-01573-y

46. Kim D, Konyn P, Sandhu KK, Dennis BB, Cheung AC, Ahmed A. Metabolic dysfunction-associated fatty liver disease is associated with increased all-cause mortality in the United States. Journal of hepatology. 2021;75:1284–1291. doi: 10.1016/j.jhep.2021.07.035

47. Varbo A, Nordestgaard BG. Directly measured vs. calculated remnant cholesterol identifies additional overlooked individuals in the general population at higher risk of myocardial infarction. European heart journal. 2021;42:4833–4843. doi: 10.1093/eurheartj/ehab293

48. Langsted A, Nordestgaard BG. Nonfasting versus fasting lipid profile for cardiovascular risk prediction. Pathology. 2019;51:131–141. doi: 10.1016/j.pathol.2018.09.062

49. Nordestgaard BG. A Test in Context: Lipid Profile, Fasting Versus Nonfasting. Journal of the American College of Cardiology. 2017;70:1637–1646. doi: 10.1016/j.jacc.2017.08.006

50. Nordestgaard BG, Langsted A, Mora S, Kolovou G, Baum H, Bruckert E, Watts GF, Sypniewska G, Wiklund O, Borén J, et al. Fasting is not routinely required for determination of a lipid profile: clinical and laboratory implications including flagging at desirable concentration cut-points-a joint consensus statement from the European Atherosclerosis Society and European Federation of Clinical Chemistry and Laboratory Medicine. European heart journal. 2016;37:1944–1958. doi: 10.1093/eurheartj/ehw152

